# Evolution of SARS-CoV-2 immune responses in nursing home residents following full dose of the Comirnaty® COVID-19 vaccine

**DOI:** 10.1101/2021.10.06.21264616

**Authors:** Estela Giménez, Juan Alberola, Ignacio Torres, Eliseo Albert, María Jesús Alcaraz, Pilar Botija, Paula Amat, María José Remigia, María José Beltrán, Celia Rodado, Dixie Huntley, Beatriz Olea, David Navarro

**Affiliations:** Microbiology Service, Clinic University Hospital, INCLIVA Health Research Institute, Valencia, Spain; Department of Microbiology, School of Medicine, University of Valencia, Valencia, Spain; Dirección de Atención Primaria, Departamento de Salud Clínico-Malvarrosa, Hospital Clínico Universitario de Valencia, Valencia, Spain; Hematology Service Clinic University Hospital, INCLIVA Health Research Institute, Valencia, Spain; Dirección de Enfermería, Departamento de Salud Clínico-Malvarrosa, Hospital Clínico Universitario de Valencia, Valencia, Spain; Comisión Departamental de control de Residencias. Departamento de Salud València Clínico Malvarrosa

**Author notes:** **Correspondence:** David Navarro, Microbiology Service, Hospital Clínico Universitario, Instituto de Investigación INCLIVA, Valencia, and Department of Microbiology, University of Valencia, Valencia, Spain. Av. Blasco Ibáñez 17, 46010 Valencia, Spain. Both authors contributed equally to the present work (Estela Giménez, Juan Alberola).

**Keywords:** SARS-CoV-2, Comirnaty®, COVID-19 vaccine, SARS-CoV-2-S antibodies, SARS-CoV-2-S T cells, Nursing home residents

## Abstract

**Objectives:** There is scarce information as to the durability of immune responses elicited by the Comirnaty® COVID-19 vaccine in nursing home residents. Here, we assessed SARS-CoV-2-Spike (S)-targeted antibody and functional T cell responses at around 6 months after complete vaccination.

**Methods:** The sample comprised 46 residents (34 females; age, 60-100 years), of whom 10 had COVID-19 prior to vaccination. Baseline (median of 17.5 days after vaccination) and follow-up (median, 195 days) plasma specimens were available for quantitation of SARS-CoV-2-S antibodies and enumeration of SARS-CoV-2-S-reactive IFN-γ CD4^+^ and CD8^+^ T cells by flow cytometry.

**Results:** In total, 44/45 participants had detectable SARS-CoV-2-S antibodies at follow-up. Overall, antibody levels were found to decrease (median, 4.8 fold). Antibodies waning was more frequent (*P*<0.001) in SARS-CoV-2 naïve (29/35) than in recovered (1/10) residents. SARS-CoV-2-S IFN-γ CD8^+^ T cells were detected in 33/46 and 24/46 at baseline and follow-up, respectively. The figures for CD4^+^ T cell counterparts were 12/46 and 30/46. Detectable SARS-CoV-2 IFN-γ CD8^+^ and CD4^+^ T cell responses at follow-up were more common in recovered (8/10 and 7/10, respectively) than in naïve residents (9/36 and 25/36, respectively). For those with detectable responses at both time points, SARS-CoV-2-S IFN-γ CD8^+^ T cell frequencies decreased significantly (*P*=0.001) over time whereas the opposite (*P*=0.01) was observed in CD4^+^ T cells.

**Conclusion:** Almost all residents displayed detectable SARS-CoV-2-S-reactive antibodies and T cell responses, respectively, by around 6 months after complete vaccination with Comirnaty® COVID-19 vaccine, albeit generally waning in magnitude over time.

## INTRODUCTION

Real-world experience has shown mRNA COVID-19 vaccines to be effective in reducing incidence of both asymptomatic and symptomatic SARS-CoV-2 infections and related deaths in nursing home residents [1], congruent with their ability to elicit robust virus-specific T and B cell immune responses in this population group [2-4]. Nevertheless, maintaining seemingly protective immune responses in these individuals over time may be compromised by the concurrence of older age, frailty and co-morbidities. To shed light on this issue, here we assessed SARS-CoV-2-Spike (S)-targeted antibody and functional T cell responses at around 6 months after vaccination with Comirnaty® (Pfizer–BioNTech) in a previously recruited cohort [2].

## MATERIAL AND METHODS

### Participants and study design

Out of 53 nursing home residents enrolled in a previous study [2] with data on B and T cell immunity at a median of 17.5 days (range, 14−35 days) after second vaccine dose (baseline sample), 46 (44 females; median age, 89 years; range, 60−100; Supplementary Table 1) were reassessed (follow-up sample) at a median of 195 days (range, 179−195 days). The remaining 7 patients either died (n=4; in no case attributable to COVID-19) or lacked the follow-up specimen (n=3). Blood specimens were collected in sodium heparin tubes (Beckton Dickinson, U.K. Ltd., UK). Informed consent was obtained from participants. The study was approved by the Hospital Clínico Universitario INCLIVA Research Ethics Committee (February, 2021).

### Immunological assays

Total antibodies (IgG and IgM) against SARS-CoV-2-S protein receptor binding domain (RBD) and the nucleoprotein (N) were measured by Roche Elecsys® electrochemiluminescence sandwich immunoassays (Roche Diagnostics, Pleasanton, CA, USA). Cryopreserved plasma (−20 ºC) specimens were thawed and assayed in singlets within 15 days after collection. Plasma specimens were diluted (1/10) for antibody quantitation when appropriate. SARS□CoV□2□S-reactive IFNγ□producing□CD8^+^ and CD4^+^ T cells were enumerated in whole blood by flow cytometry for ICS (BD Fastimmune, BD□Beckton Dickinson and Company□Biosciences, San Jose, CA) as previously described [2].

### Statistical methods

Differences between medians were compared using the Mann–Whitney U-test or Wilcoxon test for unpaired and paired data, when appropriate. The Spearman rank test was used for correlation analyses between continuous variables. Two-sided exact *P*-values were reported. A *P*-value <0.05 was considered statistically significant. The analyses were performed using SPSS version 20.0 (SPSS, Chicago, IL, USA).

## RESULTS

### SARS-CoV-2 infectious status of participants

Of the 46 residents, 10 (21.7%) had evidence of SARS-CoV-2 infection at baseline, as determined by both RT-PCR on nasopharyngeal specimens and detection of N-specific antibodies. No additional residents developed N-specific antibodies between sampling times.

### SARS-CoV-2-S-specific antibodies

Data on SARS-CoV-2-RBD antibody levels were available for 45 participants. All 43 residents who tested positive at baseline also displayed detectable responses at follow-up, although overall, antibody levels were found to decrease significantly, by a median of 4.8 fold (range, 1.1−39) [median of 2,249 IU/ml at baseline vs. median 307 IU/ml at follow-up, *P*<0.001 (Figure 1A)]. One of the two remaining residents developed SARS-CoV-2-S-specific antibodies (8 IU/ml) between sampling times. Antibodies waning was documented more frequently (*P*<0.001) in SARS-CoV-2 naïve (29/35) than in recovered (1/10) residents (Figure 1B).

**Figure 1.**
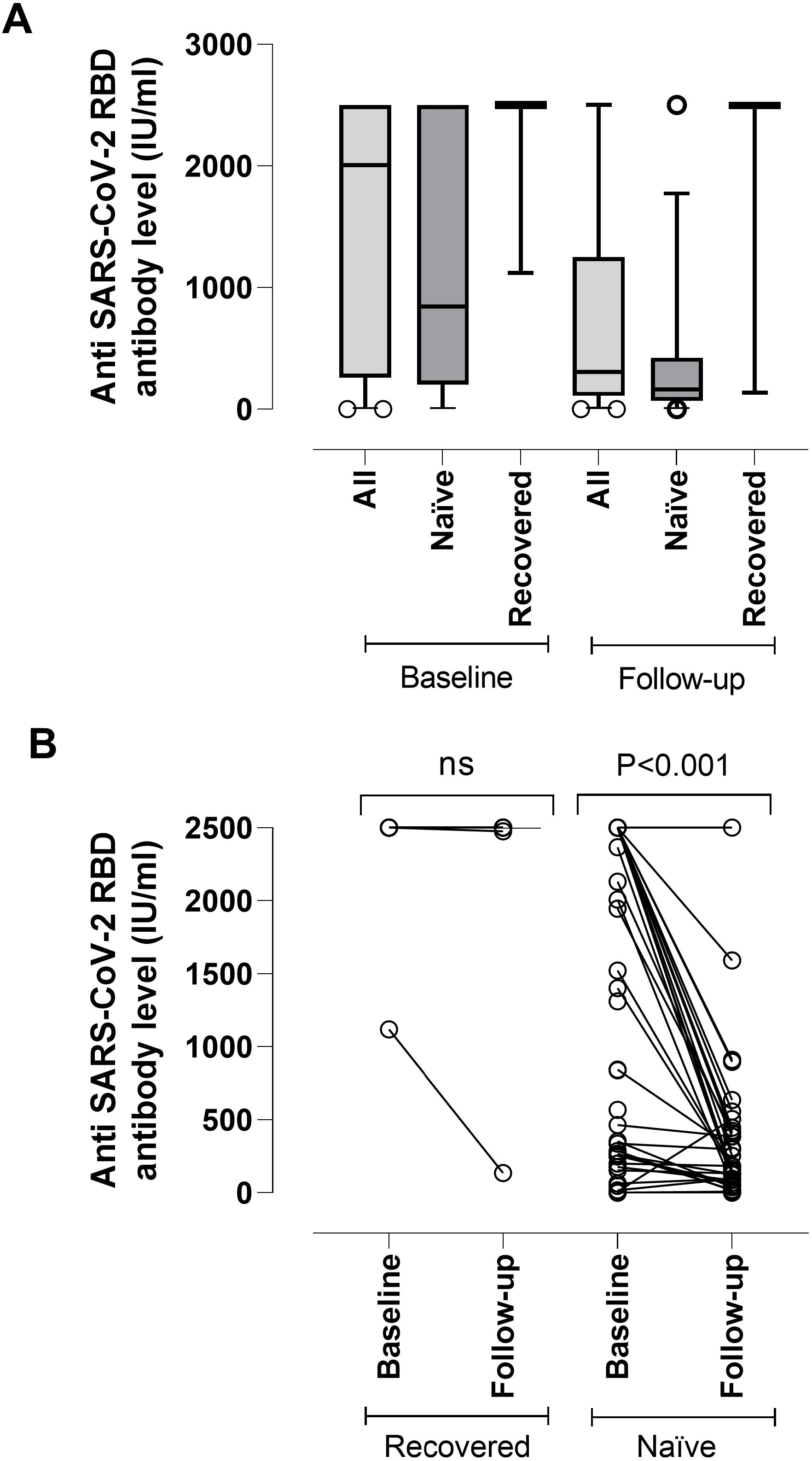
(A) SARS-CoV-2-S (RBD) plasma antibody (IgG and IgM) levels as measured by Roche Elecsys® Anti-SARS-CoV-2-S immunoassay in nursing home residents with (recovered) or without (naïve) documented prior SARS-CoV-2 infection at baseline (median, 17.5 days) and follow-up (median, 195 days) after complete Comirnaty® COVID-19 vaccination. The limit of detection of the assay is 0.4 IU/ml and its quantification range is between 0.8 and 250 IU/ml. Plasma specimens were further diluted (1/10) for antibody quantitation when appropriate. The assay is calibrated with the first WHO International Standard and Reference Panel for anti-SARS-CoV-2 antibody [12]. Bars represent median levels. (B) Individual kinetics of SARS-CoV-2-S (RBD) plasma antibodies in recovered and naïve nursing home residents. *P*-values for comparisons are shown (ns; not significant).

### SARS-CoV-2-S-specific T cells

Data on T cell responses were available for 46 participants. Overall, detectable SARS-CoV-2-S IFN-γ T cells (either CD8^+^, CD4^+^ or both) were documented in 82.6 % (38/46) and 73.9% (34/46) of residents at baseline and follow-up, respectively (*P*=0.01). The corresponding figures for SARS-CoV-2-S IFN-γ CD8^+^ T cells were 72% (33/46) and 52.1% (24/46). As shown in Figure 2A, 8 of 13 residents testing negative at baseline later acquired detectable responses, albeit at low frequencies (median, 0.08%; range, 0.01−0.21%), whereas SARS-CoV-2-S IFN-γ CD8^+^ T cells were no longer detectable at follow-up in 16 out of 33 residents who tested positive at baseline. SARS-CoV-2-S IFN-γ CD4^+^ T cells were detected in 26% (12/46) and 65.2% (30/46) of residents at baseline and follow-up, respectively. Nineteen participants developed CD4^+^ T cell responses between testing time points (median, 0.1%; range, 0.03-1.14%), whereas one out of 12 with detectable responses at baseline had lost this at follow-up (Figure 2B). The likelihood of having detectable SARS-CoV-2 IFN-γ CD8^+^ and CD4^+^ T at follow-up was higher (*P*=0.03 and *P*=0.5) in SARS-CoV-2 recovered (8/10 and 7/10, respectively) than in naïve residents (9/36 and 25/36, respectively). For those with detectable responses at both time points, overall, SARS-CoV-2-S IFN-γ CD8^+^ T cell frequencies decreased significantly (*P*=0.001) over time whereas the opposite (*P*=0.01) was seen for CD4^+^ T cells (Figure 2C). Interestingly, the resident lacking anti-RBD antibodies at follow-up had detectable SARS-CoV-2-S CD4^+^ T cell responses.

**Figure 2.**
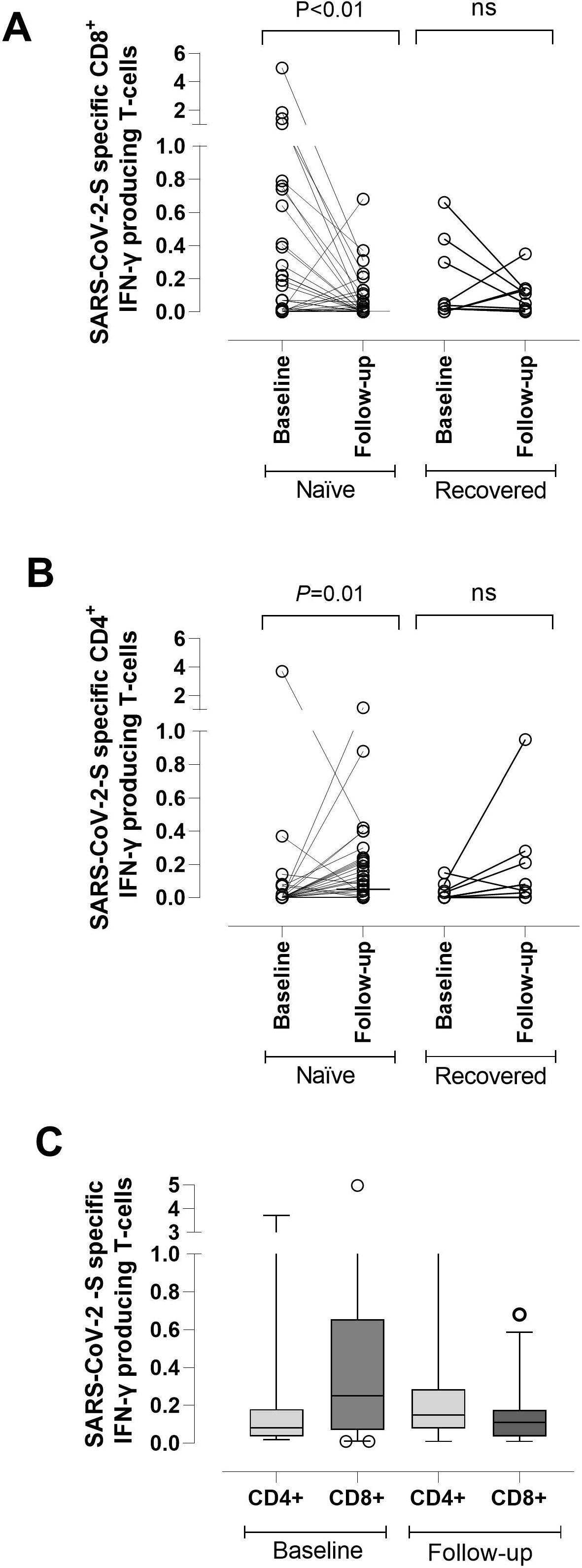
SARS-CoV-2-S-reactive IFN-γ-producing T cell levels in SARS-CoV-2 naïve and or recovered nursing home residents measured at baseline (median, 17.5 days) and follow-up (median, 195 days) after complete Comirnaty® COVID-19 vaccination. Briefly, heparinized whole blood (0.5□ml) was simultaneously stimulated for 6□h with two sets of 15□mer overlapping peptides (11□mer overlap) encompassing the SARS□CoV□2 Spike (S) glycoprotein (S1, 158 peptides and S2, 157 peptides) at a concentration of 1□μg/ml per peptide, in the presence of 1□μg/ml of costimulatory monoclonal antibodies (mAbs) to CD28 and CD49d. Peptide mixes were obtained from JPT peptide Technologies GmbH (Berlin, Germany). Samples mock-stimulated with phosphate□buffered saline (PBS)/dimethyl sulfoxide and costimulatory antibodies were run in parallel. Brefeldin A (10□μg/ml) was added for the last 4□h of incubation. Blood was then lysed (BD FACS lysing solution) and frozen at −80°C until tested. On the day of testing, stimulated blood was thawed at 37°C, washed, permeabilized (BD permeabilizing solution) and stained with a combination of labeled mAbs (anti□IFNγ□FITC, anti□CD4□PE, anti□CD8□PerCP□Cy5.5, and anti□CD3□APC) for 1□h at room temperature. Appropriate positive (phytohemagglutinin) and isotype controls were used. Cells were then washed, resuspended in 200□μL of 1% paraformaldehyde in PBS, and analyzed within 2□h on an FACSCanto flow cytometer using DIVA v8 software (BD Biosciences Immunocytometry Systems, San Jose, CA). CD3^+^/CD8^+^ or CD3^+^/CD4^+^ events were gated and then analyzed for IFN□γ production. All data were corrected for background IFN-γ production (FITC-labelled isotype control antibody) and expressed as a percentage of total CD8^+^ or CD4^+^ T cells. (A) Individual kinetics for SARS-CoV-2-S-reactive IFN-γ CD8^+^ T cells. (B) Individual kinetics for SARS-CoV-2-S-reactive IFN-γ CD4^+^ T cells. (C) Frequencies of SARS-CoV-2-S-reactive IFN-γ CD8^+^ and CD4+ T cells in nursing home residents with detectable responses at both baseline and follow-up. T cells. *P*-values for comparisons are shown (ns; not significant).

Supplementary Table 2 shows the combined results for all immunological parameters. No correlation was found between anti-RBD antibody levels and SARS-CoV-2-S IFN-γ CD4^+^ (Rho=-0.015; *P*=0.94) and CD8^+^ (Rho:-0.18; *P*=0.87) T cells.

## DISCUSSION

The main findings of the current study are as follows. First, while SARS-CoV-2-RBD antibodies, which correlate strongly with neutralizing antibody titers [5], were detectable in 97.7% (44/45) of nursing home residents by around 6 months after full vaccination with Comirnaty®, their levels declined significantly over time (a median of 5-fold); second, RBD-reactive antibody waning was rather frequent (29/35) in SARS-CoV-2 naïve, but uncommon (1/10) in recovered residents, who overall maintained high antibody levels at follow-up. The above observations were not unexpected as they have also been made in other population groups, including younger individuals seemingly with few or no comorbidities [6-8], at comparable timeframes [7,8] after full vaccination with mRNA vaccines; third, a large percentage of residents (34/46; 73.9%) had detectable SARS-CoV-2-S IFN-γ T cells (either CD8^+^, CD4^+^ or both) at follow-up, SARS-CoV-2-S IFN-γ CD4^+^ T cell responses being more frequently documented than their CD8^+^ T cells counterparts. Interestingly, recovered COVID-19 residents were more likely to display both CD8^+^ and CD4^+^ detectable responses at follow-up than naïve ones. Finally, for those with detectable responses at both sampling times, SARS-CoV-2-S IFN-γ CD8^+^ T cell frequencies decreased significantly, whereas the opposite was observed for CD4^+^ T cells. In this regard, collectively, the above data suggested that SARS-CoV-2-S IFN-γ CD4^+^ T cells may develop later than CD8^+^ T cells in nursing home residents.

Limitations of the current study are the relatively small sample size and lack of a control group; regarding the latter, most of the 17 controls included in our previous study [2] were unfortunately not available for follow-up sampling. Secondly, neutralization assays were not carried out. In summary, our data revealed that a large percentage of nursing home residents displayed detectable SARS-CoV-2-S-reactive antibodies and T cell responses, respectively, by around 6 months after complete vaccination with Comirnaty® COVID-19 vaccine, although these generally declined over time. Whether these mid-term immune responses suffice to prevent COVID-19 remains to be determined. Our data also suggested that a booster (third) dose, which has been proposed for elderly people [9,10] may be delayed beyond 6 months in fully vaccinated COVID-19 recovered residents.

## Data Availability

The data that support the findings of this study are available from the corresponding author, [DN], upon reasonable request.

## ACKNOWLEDGMENTS

We are grateful to all personnel who work at nursing home residences affiliated to the Clínico-Malvarrosa Health Department and at Clinic University Hospital, in particular to those at the Microbiology laboratory, for their commitment in the fight against COVID-19. Ignacio Torres holds a Río Hortega Contract (CM20/00090) from the Carlos III Health Institute. Eliseo Albert holds a Juan Rodés Contract (JR20/00011) from the Carlos III Health Institute. Estela Giménez holds a Juan Rodés Contract (JR18/00053) from the Carlos III Health Institute.

## AUTHOR CONTRIBUTIONS

EG, JA, IT, EA, MJA, PA, MJR, DH and BO: Methodology and data validation. PB, MJB, CR: in charge of implementing public health policies to combat SARS-CoV-2 epidemic at nursing home residences affiliated to the Clínico-Malvarrosa Health Department. DN: Conceptualization, supervision, writing the original draft. All authors reviewed the original draft.

