## Supplementary Table 1 for "Evolution of SARS-CoV-2 immune responses in nursing home residents following full dose of the Comirnaty® COVID-19 vaccine"

| **Supplementary Table 1. Characteristics of nursing home residents included in the study** | |
| --- | --- |
| **Characteristic** | **Number (%)** |
| **Sex** |  |
| Male | 12 (26%) |
| Female | 34 (74%) |
| **Comorbidities (≥1)** | 42 (91%) |
| **Comorbidities** |  |
| Hypertension | 34 (73.9%) |
| Dyslipidemia | 30 (65.2%) |
| Chronic heart disease | 17 (36.9%) |
| Diabetes mellitus | 15 (32.6%) |
| Vascular disease | 13 (28.3%) |
| Cancer | 7 (15.2%) |
| Chronic renal disease | 8 (17.4%) |
| Chronic respiratory disease | 8 (17.4%) |
| Hyperuricemia | 7 (15.2%) |
| Thyroid involvement | 7 (15.2%) |
| Obesity | 1 (2.2%) |
