## Supplementary Table 2 for "Evolution of SARS-CoV-2 immune responses in nursing home residents following full dose of the Comirnaty® COVID-19 vaccine"

| **Supplementary Table 2. Combined data on SARS-CoV-2-S- B and T-cell immunity in nursing home residents** | | |
| --- | --- | --- |
| **Qualitative results of SARS-CoV-2-S- B and T-cell immunity analyses** | **Time after complete vaccination** | |
|  | Baseline  (n=46). No. (%) | Follow-up  (n=45). No. (%) |
| Detectable CD8^+^/CD4^+^/antibodies | 6 (13%) | 18 (40%) |
| Detectable CD8^+^/antibodies | 23 (50%) | 4 (8.9%) |
| Detectable CD4^+^/antibodies | 5 (10.9%) | 9 (20%) |
| Detectable CD8^+^ | 2 (4.3%) | - |
| Detectable antibodies | 8 (17.4%) | 13 (28.9%) |
| Detectable CD4^+^ | - | 1 (2.2%) |
| Detectable CD8^+^/CD4^+^/no data on antibodies | 1 (2.2%) | - |
| Detectable CD8^+^/no data on antibodies | 1 (2.2%) | - |
